# Prehospital management of convulsive status epilepticus in children: a knowledge, attitude and practice survey of UK ambulance service clinicians

**DOI:** 10.64898/2025.12.06.25341753

**Authors:** Zakariya Vansoh, Rachael Fothergill, Fiona Bell, Ria Osborne, Andy Rosser, Caitlin Wilson, Richard F. Chin, the National Ambulance Research Steering Group

## Abstract

**Purpose:** Childhood convulsive status epilepticus (CSE) is a time-critical emergency (incidence 17-23/100,000/yr) requiring prompt treatment to reduce morbidity and mortality. Evidence shows that prehospital midazolam is as safe as diazepam, but more effective. However, legal and logistical barriers limit its use by paramedics. We performed a UK-wide survey of current practice, perceived barriers, and views on intramuscular (IM) midazolam to inform service development and research.

**Methods:** We conducted a cross-sectional JISC Knowledge-Attitude-Practice survey of personnel across all 13 UK NHS Ambulance Services (21 May–30 June 2025). The survey captured demographics, first-line antiseizure medication (ASM) choices, operational challenges, knowledge of IM midazolam, and support for research.

**Results:** 153 respondents across all 13 ambulance services (range 4–26/service) participated; 143 (93%) were paramedics (43 advanced/specialist; 99 generalist). Diazepam (rectal or intravenous (IV)) was first-line ASM in most services; buccal midazolam in three, and IM midazolam in one Trust. 89% of responders reported that alternative ASMs should be available to generalist paramedics. 97% supported research on IM midazolam for emergency treatment of childhood CSE. If approved, 85% of respondents thought their Trust would likely support the clinical use of IM midazolam. Potential barriers to IM midazolam use included dosing uncertainty (53%), risk of respiratory depression (46%), inappropriate use (37%), and reluctance to give IM injections (24%).

**Conclusion:** UK ambulance clinicians report wide variation in drug administration for prehospital seizure management, challenges with existing treatments, and support for research on paramedic-delivered IM midazolam for childhood CSE.

## Introduction

Convulsive status epilepticus (CSE) in children is a common and time-critical emergency (incidence 17 - 23 /100,000 per year) requiring prompt treatment to reduce morbidity and mortality (1, 2). Most CSE episodes start out of hospital, and half of the children with CSE have no previous neurological problems (1). Therefore, optimising prehospital treatment through paramedic services is paramount (2). Benzodiazepines are the most common first-line emergency treatment of seizures in children (3). However, it remains uncertain which type of benzodiazepine and what routes are best for paramedic delivery (4).

Obtaining intravenous (IV) access in a child with CSE prehospital can be challenging, so parenteral treatments that do not require IV access are often preferred (3). Although there is good evidence that midazolam is more effective than diazepam in stopping seizures in children (5), its use can be constrained by legal and operational factors. For example, most ambulance clinicians are not independent prescribers and therefore administer medicines under frameworks such as Patient Group Directions (PGDs) or Patient Specific Directions (PSDs), which can influence which formulations and routes are available across different services (6). Logistic issues, for example, provision and storage of prefilled syringes or ampoules and associated training, also affect practice. To our knowledge, there are limited randomized controlled trials on paramedic-led emergency treatment of seizures in children (5, 7, 8).

This survey of UK NHS ambulance service clinicians aimed to understand current practices in paramedic treatment of seizures in children, identify treatment challenges, and document views on IM midazolam to inform service development and future research.

## Methods

We carried out a cross-sectional, anonymized Knowledge, Attitude and Practice survey of key personnel in each of all thirteen UK NHS Ambulance Service Trusts in collaboration with the National Ambulance Research Steering Group. The Association of Ambulance Chief Executives (AACE) and the National Ambulance Services Medical Directors were aware and supportive of the survey. The survey ran from the 21^st^ of May 2025 – 30^th^ of June 2025. Each ambulance Trust determined which personnel within their Trust were sent the survey.

The survey consisted of questions on demographics, first-line treatments currently in use, perceived challenges and limitations of current practice, need for alternative treatment options, knowledge and awareness of intramuscular (IM) midazolam as an emergency treatment, and to gauge support and interest for research on IM midazolam as a paramedic-delivered prehospital treatment for seizures in children. (See supplemental material 1 for the survey questions).

Descriptive analyses of quantitative data were carried out in JISC Online Surveys, and supplementary analyses were performed in Microsoft Excel.

The survey was given a favorable opinion by the Edinburgh Medical School Research Ethics Committee (25-EMREC-036).

## Results

There were 153 respondents from all 13 UK NHS ambulance services; respondents per service ranged from 4–26. 143 (93%) were paramedics, of which 43 (28%) were chief/advanced/specialist and 99 (65%) were generalist paramedics. The remaining respondents were one doctor, two nurses, three non-registered ambulance staff, one pharmacist, and “others”.

Diazepam by the rectal or intravenous route is first line Anti-Seizure Medication (ASM) for childhood CSE in most UK ambulance services, three Trusts use buccal midazolam as first-line ASM, and one Trust uses IM midazolam. Overall, 89% of respondents think there is a need for alternative ASMs to be made available for generalist paramedics for the treatment of children with seizures. In Trusts that use diazepam as first-line ASM, a higher percentage of respondents thought alternative ASM was needed compared to respondents from Trusts that are currently using midazolam (94% vs 75%, chi-square 10.3, p=0.001).

One third of respondents either currently use IM midazolam in their service or are aware of its use in other places. These include outside of the UK (New Zealand, Australia, the Republic of Ireland, and Jersey) and within the UK (mainly London and in emergency departments). If approved for clinical use, 85% of respondents thought their service would support IM midazolam in their Trust; there was no difference in the proportion expressing support between respondents from Trusts that already use buccal or IM midazolam and those from Trusts that use diazepam as first line (89% vs 83%, p=0.45).

The main challenges in current practice reported by paramedics are obtaining IV access in children having seizures, resistance to administering medication rectally, and training in the identification of seizures.

Most reported perceptions of potential challenges to the use of IM midazolam by paramedics were uncertainty about dosage (53%), risk of respiratory depression (46%), use in patients not having seizures (37%), and reticence of administering IM medication (24%). 97% of responders were supportive of research on paramedic-delivered IM midazolam for the emergency treatment of childhood CSE.

## Discussion

The main findings of this survey are that in the UK, diazepam remains the main first-line emergency ASM for childhood CSE in ambulance services; paramedics want to have access to alternative ASMs, particularly those working in services using diazepam; and there is support for research and clinical use of IM midazolam if approved.

Since the survey, the current Joint Royal Colleges Ambulance Liaison Committee guidelines list midazolam (IM or buccal) or diazepam (by the intravenous or rectal route) as first-line ASM for childhood CSE, but with a clear steer that midazolam should be used whenever possible, given its superiority to diazepam (9). Having multiple medication options has practical advantages in that it supports care continuity during supply issues and addresses potential legal restriction. However, it also probably reflects ongoing equipoise regarding optional ambulance-based treatment for childhood CSE in the UK.

The almost unanimous support for research on IM midazolam for childhood CSE suggests there would be much interest amongst UK ambulance services for a multi-site UK wide trial. A network metanalysis of mainly paediatric trials of non-intravenous treatment of CSE found IM midazolam had a shorter time to seizure cessation, but a similar low incidence of adverse effects, compared to other treatments (10). A cost-effectiveness metanalysis of paediatric studies of non-intravenous rescue medications found IM midazolam stopped seizures in 88% (95%CI 73-95%) and 73% (95% CI 60-83%) in those treated with buccal midazolam (8).

This is the only cost effectiveness study on non-intravenous first line ASM, but this was based on USA costs, and therefore, cost-effectiveness in the UK setting is unknown. To our knowledge, there is no clinical trial in the ambulance setting for childhood CSE comparing IM midazolam with midazolam by another route, or any other benzodiazepines by any non-intravenous route.

Legally, paramedics are allowed to administer diazepam, but not midazolam, without a prescription, as the former but not the latter is included in Schedule 17 of the Human Medicines Regulations (11). A current consultation forming part of a broader review of Schedule 17 medicines to enhance emergency preparedness (12) proposes adding injectable midazolam to Schedule 17 exemptions (12). If approved, this could help pave the way for easier paramedic use of midazolam for emergency childhood CSE treatment, if there are trial data showing its benefit and safety.

This survey has limitations. It used a convenience sample, which may not represent all clinicians or services. As the survey was anonymous, participants’ follow-up to clarify responses or gather further context was not possible.

## Conclusion

There is a need to standardise paramedic treatment of seizures in children in the UK. NHS Ambulance staff are supportive of investigating IM midazolam as a potential prehospital treatment for childhood CSE.

## Supporting information

Trial Survey Text

## Data Availability

All data produced in the present study are available upon reasonable request to the authors.

