## Supplementary material for "Prehospital management of convulsive status epilepticus in children: a knowledge, attitude and practice survey of UK ambulance service clinicians": Trial Survey Text

### Prehospital first line Emergency Treatment of Seizures in Children (PETS) – a survey of UK ambulance professionals

**BACKGROUND**

Seizures lasting at least 5 minutes are more likely to go on for at least 30 minutes. Such prolonged seizures (status epilepticus) are the most common medical neurological emergency in childhood. They are associated with death and neurological and neurodevelopmental sequelae. Earlier, effective antiseizure treatment may improve outcomes.

Since most seizures in children start out of hospital, and 50% have never previously had seizures, **optimising prehospital treatment of seizures by ambulance services could improve neurological outcomes of children with seizures**. During the most recent review of the Joint Royal Colleges Ambulance Liaison Committee's guidelines for the emergency treatment of seizures in children, there were anecdotal reports of variation in practice and uncertainty about what is optimal paramedic emergency treatment of seizures in children.

This survey is aimed at systematically capturing current practice, perception on the need for additional options, and perceived challenges for use of intramuscular midazolam for the emergency treatment of seizures in children. **Results from this survey would help development of a trial of prehospital intramuscular midazolam for seizures in children**. We are seeking survey responses from several ambulance professionals, rather than only one person, per ambulance service.

Thank you in advance for completing this short survey (it should take no more than 5 minutes).

Richard Chin
Professor of Paediatric Neurology and Clinical Epidemiology
Hon Consultant Paediatric Neurologist
The University of Edinburgh
Royal Hospital for Children and Young People Edinburgh

**1.** What area of the UK are you based?
- East of England
- East Midlands, England
- London, England
- North East, England
- North West, England
- Northern Ireland
- Scotland
- South Central, England
- South East Coast, England
- South West, England
- Wales
- West Midlands, England
- Yorkshire, England
- I prefer not to answer

**2.** What is your job title?
- Medical Doctor
- Nurse
- Ambulance Service Medical Director
- Ambulance Service Associate Medical Director
- Chief/Advanced/Specialist Paramedic
- Paramedic
- Non-registered ambulance staff
- Ambulance Pharmacist

- I prefer not to answer
- Other (free text option)

**Current practice:**

**3.** If the patient does not already have a designated emergency care plan, what is the usual preferred first line anti-seizure medication (ASM) in children in your service?

- Rectal diazepam
- Buccal midazolam
- Intranasal midazolam
- Intravenous lorazepam
- Intravenous diazepam
- Intravenous midazolam
- Low dose (0.1- 0.15mg/kg) intramuscular midazolam
- High dose (0.2-0.4mg/kg) intramuscular midazolam
- I prefer not to answer
- Other

If other, please state:

**4.** If the patient does not already have a designated emergency care plan, and you are unable to give the preferred first line ASM of your service, what is the next preferred alternative?

- Rectal diazepam
- Buccal midazolam
- Intranasal midazolam
- Intravenous lorazepam
- Intravenous diazepam
- Intravenous midazolam
- Low dose (0.1- 0.15mg/kg) intramuscular midazolam
- High dose (0.2-0.4mg/kg) intramuscular midazolam
- I prefer not to answer
- Other

If other, please state:

**5.** If the patient does not already have a designated emergency care plan, aside from buccal midazolam or rectal diazepam, what other emergency first line ASMs do you have access/permission to give in these situations? Choose as many as applicable.

- Intranasal midazolam
- Intravenous lorazepam
- Intravenous diazepam
- Intravenous midazolam
- Low dose (0.1- 0.15mg/kg) intramuscular midazolam
- High dose (0.2-0.4mg/kg) intramuscular midazolam
- None
- I prefer not to answer
- Other

If other, please state:

**6.** Do you think that there is a need for alternative ASMs being made available for standard (ie not advanced/specialist) paramedics for treatment of children with seizures?

- Yes
- No
- DK
- I prefer not to answer

**7.** What do you think are the perceived challenges/limitations of current practice? (Free text)

**8.** If intramuscular midazolam is not a part of your local guideline for emergency treatment of seizures in children, are you aware of im midazolam being used for emergency treatment of seizures in children in other places?

- Yes
- If yes, please tell us where
- No
- DK
- I prefer not to answer

**9.** Do you think your service would support the clinical use of intramuscular midazolam by pre-hospital clinicians?

- Yes
- No
- DK
- I prefer not to answer

**10.** What would you perceive as potential challenges to use of im midazolam by prehospital clinicians? Choose as many as applicable.

- Inappropriate use ie treatment when patient not having seizures
- Use by non-registered ambulance staff rather than paramedics
- Reticence of administering intramuscular medication
- Worry about local complications eg bleeding, infection
- Concerns about risk of respiratory depression
- Uncertainty about dosage
- Lack of effectiveness
- I prefer not to answer
- Other

If other, please state:

**11.** Would you be supportive of research to explore the use of im midazolam by paramedics for children with seizures?

- Yes
- No
- DK
- I prefer not to answer

**12.** Please indicate below how many patients in your service might be eligible in one year for a proposed study of paramedic emergency treatment of seizures in children. If you have already submitted this information to Professor Richard Chin, please ignore the question. If you do not have this information readily available, and need to carry out an audit/data query, please feel free to email your results when they are available to Richard Chin as above.

NB! This information on number of potential research participants will be needed when advertising the study and for selection of trial sites.
